# Monitoring peripheral hemodynamic response to changes in blood pressure via photoacoustic imaging

**DOI:** 10.1101/2022.02.23.22271420

**Authors:** Yash Mantri, Tyler R. Dorobek, Jason Tsujimoto, William F. Penny, Pranav S. Garimella, Jesse V. Jokerst

## Abstract

Chronic wounds and amputations are common in chronic kidney disease patients needing hemodialysis (HD). HD is often complicated by drops in blood pressure (BP) called intra-dialytic hypotension. Whether intra-dialytic hypotension is associated with detectable changes in foot perfusion, a risk factor for wound formation and impaired healing remains unknown. Photoacoustic (PA) imaging is ideally suited to study perfusion changes. We scanned the feet of 20 HD and 11 healthy subjects. HD patients were scanned before and after a dialysis session whereas healthy subjects were scanned twice at rest and once after a 10 min exercise period while BP was elevated. Healthy (r=0.70, p<0.0001) and HD subjects (r=0.43, p<0.01) showed a significant correlation between PA intensity and systolic BP. Furthermore, HD cohort showed a significantly reduced PA response to changes in BP compared to the healthy controls (p<0.0001). Hence showing that PA can monitor hemodynamic changes due to changes in BP.

## 1. Introduction

Chronic kidney disease (CKD) affects more than 9% of the global population.[1, 2] An estimated two million of these patients with CKD progress to end-stage kidney disease (ESKD) and must undergo kidney replacement therapies such as a kidney transplant or dialysis.[1] Rates of non-traumatic lower limb amputations are ∼4.3/100 person-years for ESKD patients and reach 13.8/100 person-years for the diabetic subpopulation.[3] Foot ulcers usually precede 84% of amputations with half occurring in diabetic patients.[4] Worse, diabetic dialysis patients develop foot ulcers at five-fold higher rate than even diabetic chronic kidney disease patients.[5] Foot ulceration is a significant risk factor for limb loss, and thus prevention—along with timely diagnosis and treatment—may translate to a reduced amputation rate. There is an urgent need to develop novel, non-invasive techniques to diagnose risk factors for ulceration and prevent limb loss in persons with ESKD.

Clinically, tissue perfusion is often inferred using measurements of blood pressure (BP),[6] blood oxygen saturation,[7] and lactate levels.[8] Measurements of full body hemodynamic parameters fail to reflect changes in peripheral microcirculation.[9] Transcutaneous oxygen monitoring (TcOM),[10] functional magnetic resonance imaging (fMRI),[11] laser doppler imaging (LDI)[12], and spatial frequency domain imaging (SFDI)[13] can measure local perfusion and oxygenation in specialized care settings. TcOM is a non-invasive skin oxygen tension measurement system used for infants and adults.[10] But TcOM suffers from long acquisition times (15-20 min) and is susceptible to calibration errors and poor inter-rater usability.[14] fMRI can non-invasively image difficult to access areas like the brain and produce perfusion maps.[11] But fMRI has poor temporal resolution (5 sec) and reproducibility.[15, 16] LDI can provide real-time and continuous perfusion monitoring but is sensitive to all movements resulting in erroneous readings, and it can only penetrate a few millimeters into tissue.[17] SFDI can provide a large field of view and rapid imaging but is limited to surface tissues and still needs to be developed further for clinical use.[13]

Photoacoustic (PA) imaging is a hybrid imaging modality that can solve these major limitations. PA imaging uses pulsed light to generate sound waves via thermal expansion that can be overlaid with conventional ultrasound (US) data.[18] PA employs the difference in light absorption between oxygenated and deoxygenated hemoglobin to measure oxygen saturation, map vasculature and tissue perfusion in real time.[19]

In pre-clinical settings, PA is used to measure disease biomarkers,[20] oxidative stress,[21, 22] blood oxygen saturation,[23] and image chronic wounds.[24, 25] Clinically PA is used for diagnosing and monitoring breast cancer progression, [26, 27] vascular dynamics in human fingers, [28] inflammatory bowel disease, [29] and many more diseased states.[30-33] Recently, we demonstrated the ability of PA imaging to monitor angiogenesis and predict wound healing.[34] The non-invasive, real-time, and enhanced penetration depth (>4 cm) of PA imaging make it an ideal tool to map peripheral tissue perfusion as a risk factor for the development of limb complications.

Here, we aimed to explore the use of PA to measure changes in peripheral blood perfusion in a cohort of HD patients before and after dialysis. We hypothesized that the change in PA signal would be dependent on changes in blood pressure given the intradialytic hypotension that often occurs in patients.[35-39] The loss of pressure is followed by peripheral vasoconstriction and reduced peripheral perfusion because blood flow is redirected towards the vital organs.[40] This controlled change in blood pressure in a resting patient without medication or exercise is unique to HD patients. Hence HD patients make an exquisite cohort to study the effects of blood pressure on peripheral photoacoustic signals and tissue perfusion.

## 2. Methods

### 2.1 Patients

This study was performed in accordance with the ethical guidelines for human experimentation stated in the 1975 Declaration of Helsinki. The study was approved by the University of California San Diego’s Human Research Protections Program and was given Institutional Review Board approval (IRB# 191998). Written informed consent was obtained from all subjects before participation. All subjects were ≥18 years old and able to provide consent. Those in the CKD group were all on hemodialysis. Exclusion criteria were: (i) presence of bloodborne pathogens and (ii) presence of implants in the imaging region. Dialysis patients (n=22) were recruited consecutively for this study at the outpatient Hemodialysis Unit, UC San Diego Health System. Eleven healthy volunteers, with no known vascular disease history were recruited at UC San Diego.

The dialysis patients were each scanned twice: once at the start of HD (pre-dialysis), and again at the completion of HD (post-dialysis). Blood pressure and ultrafiltration volume were recorded by the Fresenius 2008T dialyzer (Fresenius Medical Care, Waltham, MA, USA). Two of the 22 dialysis patients were subsequently excluded from the analysis due to complications during HD (unrelated to imaging) preventing a post HD scan.

The 11 healthy subjects were each scanned three times: 1) at baseline; 2) after 3 hours of rest and no pressure changes (negative control (test/re-test), pre-exercise); and 3) after 10 minutes of exercise to elevate blood pressure (positive control, post-exercise). We monitored body temperature, heart rate, and blood pressure at all imaging time points. **Fig. 1** illustrates the study protocol for the two groups. **Table 1** describes the CKD/HD and healthy group demographics.

**Figure 1.**
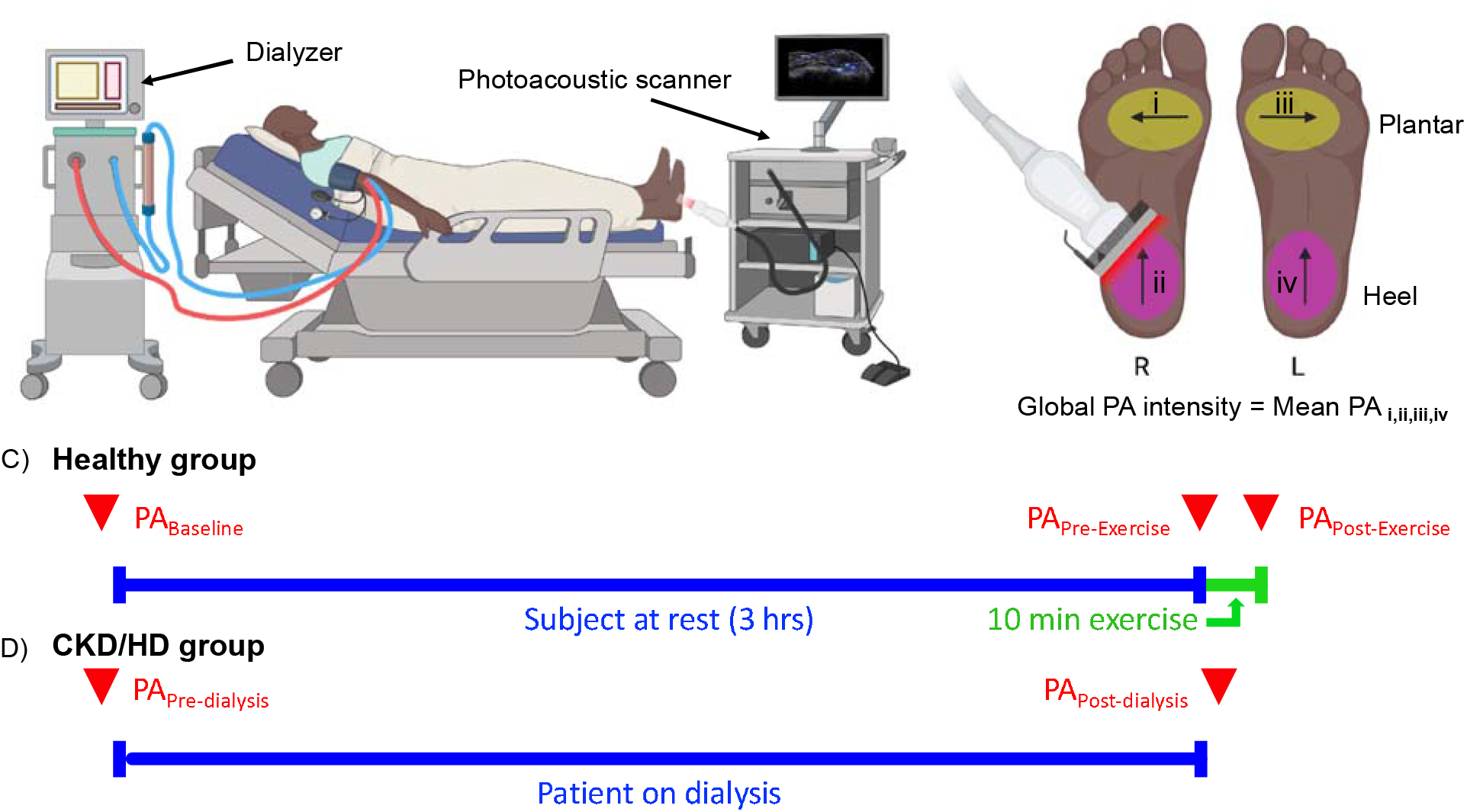
Photoacoustic monitoring of peripheral perfusion; study design and timeline. **A**. The CKD/HD group consisted of patients on hemodialysis (HD). **B**. We scanned the plantar and heel area in a medial-lateral and inferior-superior direction, respectively. Global PA intensity was defined as the mean PA intensity of all four imaging regions. **C**. Healthy subjects were scanned at baseline (T = 0 hrs), pre-exercise (T = 3 hrs), and immediately after 10 min of exercise (post-exercise). **D**. HD patients were scanned pre-and-post their HD session. Red downward triangles represent imaging time points. Blue line indicated healthy subjects at rest for 3 hrs and CKD/HD patients on dialysis. Green denotes the 10 min exercise period to increase blood pressure in healthy subjects.

**Table 1.**
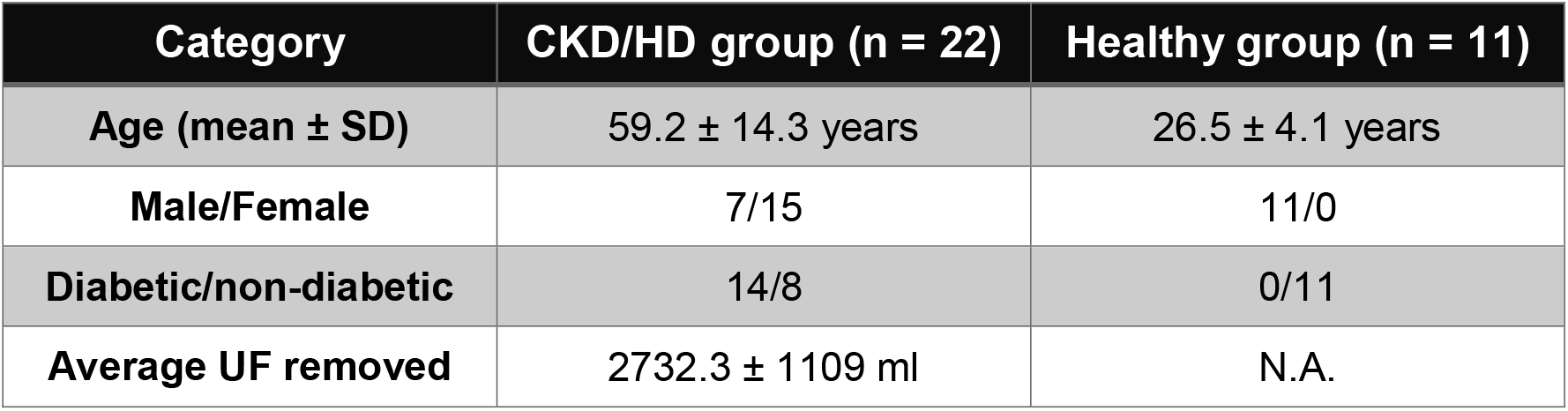
CKD/HD and Healthy group demographics.

| Category | CKD/HD group (n = 22) | Healthy group (n = 11) |
| --- | --- | --- |
| Age (mean ± SD) | 59.2 ± 14.3 years | 26.5 ± 4.1 years |
| Male/Female | 7/15 | 11/0 |
| Diabetic/non-diabetic | 14/8 | 0/11 |
| Average UF removed | 2732.3 ± 1109 ml | N.A. |

### 2.2 Blood pressure measurements

Blood pressure (BP) for the HD patients was measured internally by the dialyzer. We also noted their ultrafiltration volume at the end of the HD session. Patients were at rest sitting in semi-Fowler’s position at the time of measurement. BP measurements were made just before and at completion of the HD session (**Fig. 1D**). Change in systolic pressure was defined as the difference between BP_HDend_ and BP_HDstart_.

For the healthy group we used an electronic blood pressure monitor (Omcron Healthcare Inc., Lake Forest, IL, USA, Model no: BP742N). The cuff was placed on the left upper arm and the subjects were sitting up straight for all measurements. We recorded BP twice at rest (T=0 and T=3 hrs) and once after 10 min of exercise (**Fig. 1C**). The T=3 hrs time point was chosen to mimic the length of a typical HD session without changes in BP, serving as a negative control. Exercise consisted of climbing up and down stairs for 10 min to simulate an increase in blood pressure (positive control). Changes in systolic BP were calculated as differences between (i) BP_pre-exercise_ and BP_baseline_; and (ii) BP_post-exercise_ - BP_pre-exercise_.

### 2.3 Photoacoustic imaging

All the PA imaging was done using the AcousticX from Cyberdyne Inc. (Tsukuba, Japan). This LED-based PA scanner operates at 850 nm wavelength with an LED repetition rate of 4 kHz, pulse width of 70 ns, and operating at 2.6 µJ/cm^2^ per pulse.[41] It employs a 128-element linear array transducer with a 0.38 × 6 cm field of view, 7 MHz central frequency, and a bandwidth of 80.9%.[42] We used a custom hydrophobic gel pad from Cyberdyne Inc. along with sterile US coupling gel (Aquasonic 100, Parker Laboratories Inc. Fairfield NJ, USA) to couple the transducer with the skin surface. A sterile sleeve (CIV-Flex™ #921191 from AliMed Inc., Dedham, MA, USA) covered the transducer during imaging. All images were acquired at 30 frames/s in a single handheld sweep.

We scanned two spots on each foot (except for one patient in whom only one foot was scanned due to a left lower limb amputation). The plantar area was scanned in a medial-lateral direction and the heel area in an inferior-superior direction (**Fig. 1B**). It must be noted that since all the scans are done manually, precise control of the scan area was extremely difficult. But the same general area was scanned for every patient. CKD/HD patients were scanned once before and once after an HD session (**Fig. 1D**). Healthy subjects (the control group) were scanned three times (**Fig. 1C**). BP measurements and PA imaging were carried out simultaneously.

### 2.4 Image processing

All scans were reconstructed and visualized using the proprietary AcousticX software (Cyberdyne Inc. version 2.00.10); 8-bit PA, B-mode, and PA+US overlayed coronal cross-sectional images were exported. We used the proprietary software developed by the PA system’s manufacturer for image reconstruction. The system reconstructs PA images using Fourier transform analysis.[41] All the processing was done manually. Scans ranged between 45 – 180 frames governed by scan distance and length. PA signal was quantified using region of interest (ROI) analysis *via* ImageJ (with Fiji extension), version 2.1.0/1.53c. We drew a 4-cm-wide and 1-cm-deep rectangular ROI that was kept constant across all frames. PA signal from the skin was excluded from analysis to minimize the impact of variable melanin concentration in different skin tones. Integrated density function was used to quantify the PA signal within the ROI. Global PA intensity was defined as the mean integrated PA density of all four imaging regions (**Fig. 1B**).

### 2.5 Statistics

We measured changes in PA intensity as a function of changes in systolic BP. Simple linear regression was used to fit this data and plotted with 95% confidence intervals. We used a paired, two-tailed t-test to compare PA intensities pre-and-post exercise and HD for the healthy and diseased groups, respectively. We also used a student’s t-test to compare the hemodynamic response to changes in blood pressure for HD *vs*. healthy subjects. A p-value < 0.05 was considered significant. We tested equality of the two population variances (Healthy *vs*. CKD/HD group) using a one-tailed F-test with an alpha = 0.05. The difference was considered significant if the F-value was less than the F_critical_ value. We also ran two separate multivariate linear regressions (CKD/HD and healthy group) to study the effect of other confounding variables such as body mass index (BMI in kg/m^2^), diabetic status (Y/N), UF removed (ml), age (years), sex (M/F). A covariance matrix was also calculated to study the effect of the confounding variables on each other. Future work will evaluate heart rate and body temperature in this multivariate analysis.

## 3. Results

A total of 33 subjects were recruited for this study. All subjects were pseudonymized with an identification code at the start of the study. The healthy and the CKD/HD group are referred as HC 0XX and CKD/HD 0YY respectively. Patient demographics and BP data can be found in **Tables S1** (Healthy) and **S2** (CKD/HD).

### 3.1 Healthy control group

Eleven subjects (26.5 ± 4.1 years old) with no history of cardiovascular or other disease were recruited at UCSD. Subjects were scanned at baseline (T = 0 hrs), pre-exercise at rest (T = 3 hrs), and immediately post-exercise (**Fig. 2A**). Exercising consisted of climbing up and down stairs for 10 minutes. We monitored BP, heart rate, and body temperature at each imaging time point (**Table S1**). **Fig. 2** shows PA data from the healthy cohort.

**Figure 2.**
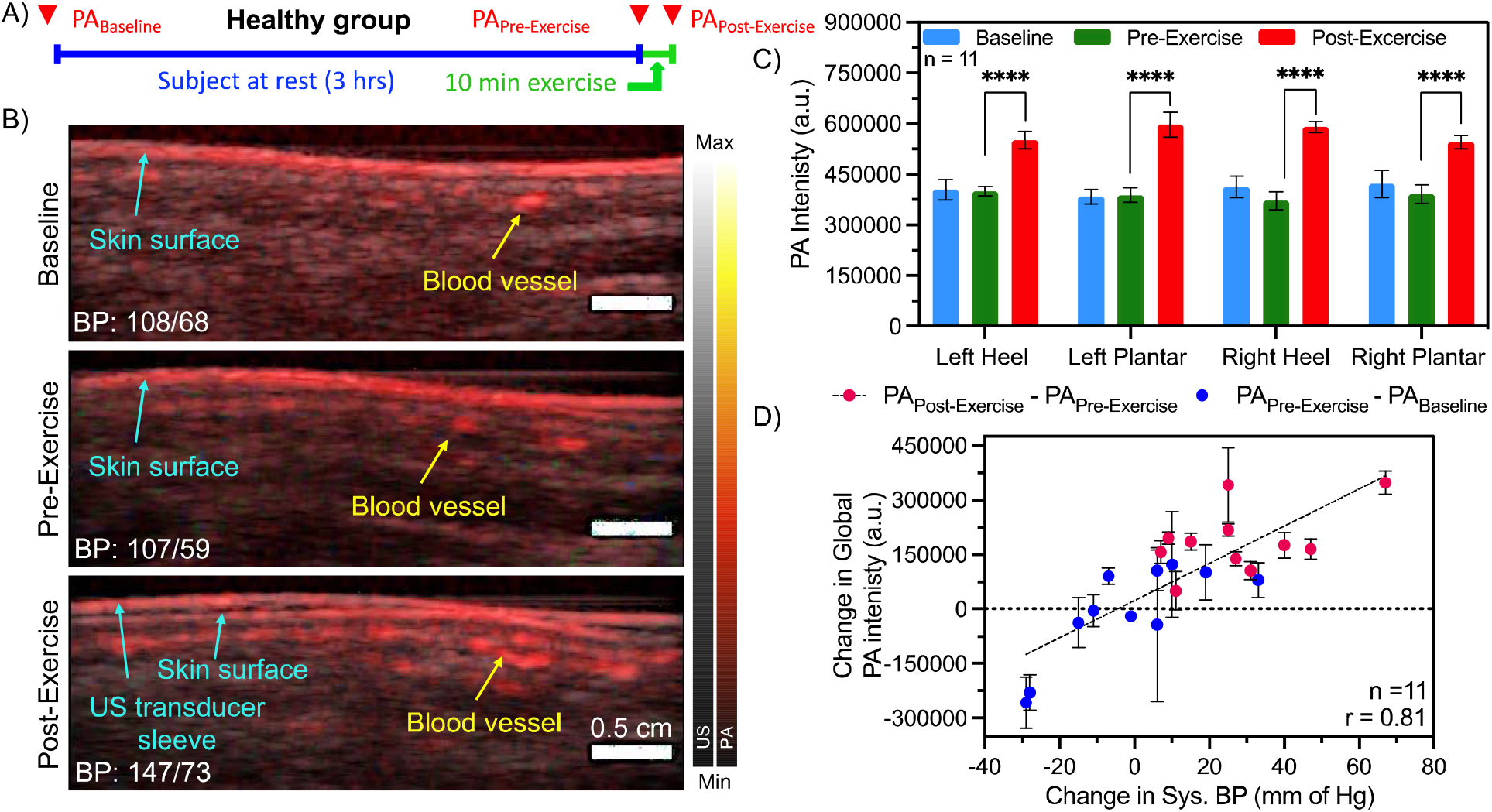
Hemodynamic response to changes in blood pressure for healthy subjects. **A**. Study design for the healthy group. Subjects were scanned at baseline (T = 0 hrs), pre-exercise at rest (T = 3 hrs), and immediately post-exercise. **B**. PA and US overlay of the left heel (HC 002). Blood vessels appear as distinct red dots marked by the yellow arrow. The skin surface is labelled in blue. At rest there was negligible change in systolic BP between baseline and pre-exercise. After a 10 min exercise session, blood pressure increased by 40 mm of Hg accompanied by a higher PA signal. PA intensity was quantified using a rectangular ROI measuring 4 cm x 1 cm. The skin surface was excluded from analysis. The ROI annotated images can be found in the supplementary information. Scale bar represents 0.5 cm. **C**. PA intensity was significantly higher after exercise (****, p<0.0001, n= 11) at each imaging site. There were no significant differences between PA_Baseline_ and PA_Pre-Exercise_. This rest period served as a negative control for the healthy group. **D**. The change in global PA intensity in the healthy group correlated with the change in systolic BP (r = 0.81, p<0.001). The 22 data points reflect changes between PA_Pre-Exercise_ – PA_Baseline_ (Blue) and PA_Post-Exercise_ – PA_Pre-Exercise_ (Pink) (n=11). Error bars in panel C represent standard deviation between 11 subjects and in panel D at least 90 frames.

The PA signal under the skin surface increased by 47% (p<0.01), and the average BP increased by 27.6 ± 18.2 mmHg immediately post-exercise relative to baseline (**Fig. 2 B-C**). The change in PA intensity was directly (r = 0.81) and significantly (p<0.0001) proportional to the change in systolic BP (**Fig. 2D**). PA intensity was significantly (p<0.0001) higher after exercise when BP was elevated (**Fig. 2C**). As a negative control, the subjects were maintained at rest for 3 hrs, simulating the length of a typical HD session. There were no significant changes in BP and PA intensity (baseline - pre-exercise) during the rest period (p = 0.18).

### 3.2 CKD/HD group

Twenty-two HD patients were enrolled in this cohort (59.2 ± 14.3 years old). Two patients developed hypotension-related complications preventing a second scan and were not included the analysis due to incomplete data acquisition (**Table S2**). The complications were independent of the imaging study. Patients were scanned before and after a routine HD session (**Fig. 3B**).

**Figure 3.**
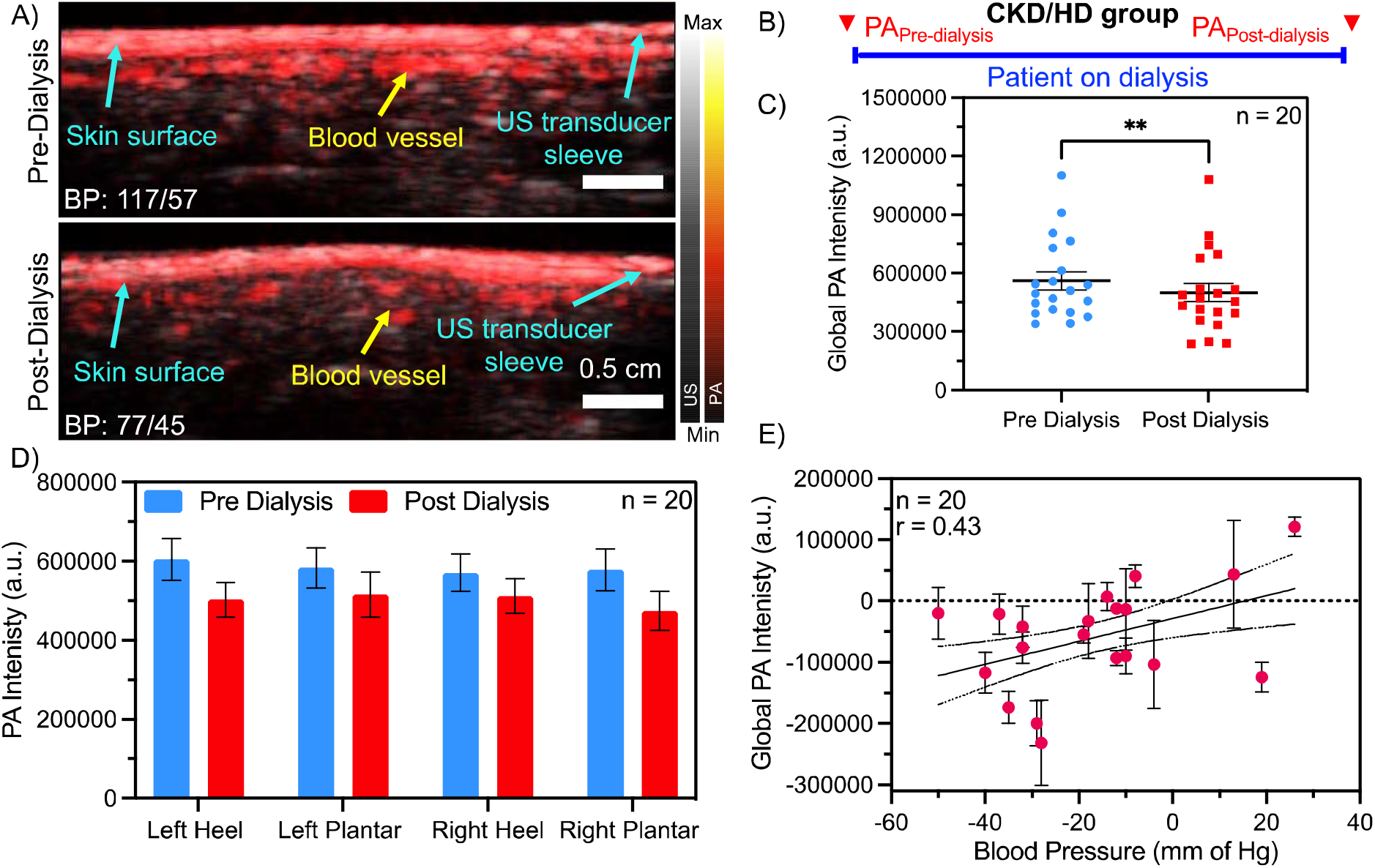
Changes in peripheral tissue perfusion during hemodialysis. **A**. PA and US overlay of the left heel in subject CKD/HD 012. Blood perfusion into the heel is considerably lower after dialysis when BP is low. Scale bar represents 0.5 cm. **B**. CKD/HD patients were scanned just before and after a routine dialysis session. **C**. A paired t-test showed a significant decrease in global PA signal and hence perfusion after dialysis (**, p<0.01). **D**. PA intensity at individual imaging areas were lower (not significantly) after dialysis. **E**. Hemodynamic response of the CKD group to changes in BP during dialysis showed a positive correlation (r = 0.43, p<0.01). Error bars in panel C-D represent standard deviation among 20 patients and in panel E at least 90 frames.

On average PA signal and systolic BP reduced by 11% and 16.6 ± 19.8 mm of Hg respectively. PA intensity and hence perfusion was significantly lower after dialysis (**Fig. 3C**, p<0.01). PA intensity in different imaging regions trended lower after dialysis but the difference was not statistically significant. PA intensity showed a positive correlation to changes in BP during an HD session (r = 0.43, p<0.01, **Fig. 3E**). PA signal and hence perfusion was higher pre-dialysis when BP was higher. The UF removed showed a positive correlation with BP but did not directly correlate with the change in PA intensity (**Fig. S4**)

### 3.3 Healthy vs. CKD/HD subjects

HD patients showed a significantly different PA response to changes in BP compared to healthy controls (**Fig. 4**). The mean PA intensity was significantly higher for the HD group (**Fig. 4A**, p<0.01). More importantly, the data spread (standard deviation) for the diseased group was significantly wider (F = 0.045; F_critical_ = 0.63). The hemodynamic response to changes in BP was characterized using the slope of changing PA as a function of changing systolic pressure (**Fig. 4B**). The slope for the HD group (1870 a.u. intensity/mm Hg) was significantly (p = 0.0001) lower than the healthy group (5116 a.u. intensity/mm Hg).

**Figure 4.**
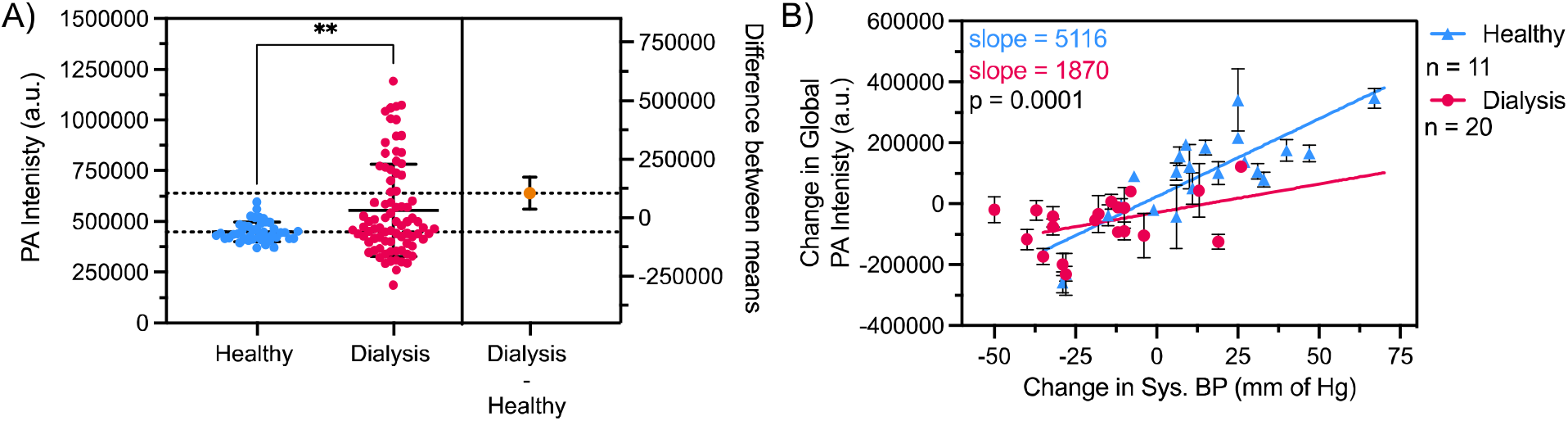
Comparing hemodynamic response to changes in BP in health *vs*. dialysis patients. **A**. PA intensities of the healthy and diseased group for all scans. CKD/HD show a significantly wider PA distribution (F = 0.045; F_critical_ = 0.63) compared to the healthy group. Dialysis patients also showed a significantly higher (**, p<0.01) mean PA intensity compared to healthy control group due to higher mean BP. **B**. Healthy subjects showed a significantly higher slope (significant difference between slopes; p = 0.0001) to changes in BP compared to the CKD/HD patients. The slope of the simple linear regression characterizes the hemodynamic response to changes in BP. Error bars in panel A represent standard deviation among all scans (n_healthy_ = 44; n_diseased_ = 80) and in panel B at least 90 frames.

## 4. Discussion

This cohort study explored the use of photoacoustic imaging to compare changes in peripheral perfusion in response to varying BP in HD patients *vs*. healthy controls. Our data suggests that HD patients show a significantly reduced PA response (p = 0.0001) to changes in BP compared to healthy controls.

The removal of excess fluid during dialysis as UF increases hematocrit.[35] Since PA leverages hemoglobin in red blood cells to generate contrast, the increase in hematocrit should increase PA intensity after dialysis.[43, 44] But we observed the opposite effect: The global PA intensity significantly decreases after HD (**Fig. 3C**, p<0.01). This can be attributed to the loss of BP after dialysis. A sudden loss in blood pressure results in peripheral vasoconstriction as blood is shunted towards the vital organs to preserve their function.[45] Vasoconstriction results in reduced peripheral perfusion that is observed as lower PA intensity after HD. Hypertension is present in 68% of HD patients, and this further reduces peripheral perfusion.[46] The changes in peripheral perfusion can exacerbate the risk of developing complications such as ulceration, amputations, and cardiovascular morbidity.[47-49]

The management of BP in HD patients is challenging but extremely important. A sudden drop in BP can leave patients lethargic and weak.[50] Some patients show a paradoxical rise in blood pressure after dialysis due to an increased cardiac output and hematocrit.[51, 52] Three HD patients (13.6%) showed a paradoxical rise in BP in the HD cohort, which might explain why individual imaging regions showed no significant difference before and after dialysis (**Fig. 3D**). Instead, a paired comparison of the global PA intensity in **Fig. 3C** showed a statistically significant difference (p<0.01).

The main clinical finding of this work is that photoacoustic imaging can be used to differentiate between a healthy and diseased response to changes in BP (**Fig. 4B**). The slopes characterize the change in perfusion as a function of BP. The healthy control cohort showed a significantly higher perfusion (p = 0.0001) when BP was elevated compared to the CKD/HD cohort. HD patients had a significantly wider distribution of PA intensities due to higher variability in BP compared to the control group (HD: 135.4 ± 29.1 mm of Hg; Healthy: 137.3 ± 19.4 mm of Hg, **Fig. 4A**). Although this cohort study was not age and gender matched, we have 85% power in our results (**Fig. S5**). Furthermore, the healthy group showed a significant positive correlation between global PA intensity and absolute systolic BP (**Fig. S6**, r = 0.70, p<0.05) at baseline. The CKD/HD group showed no significant correlation which can be attributed to varying disease etiologies and progression. Hence, this work suggests that PA imaging can be used to monitor peripheral tissue perfusion, a risk factor for wound formation and impaired healing.[53, 54] Others have also shown the use of PA imaging to visualize tissue perfusion but most studies use high-powered lasers which are expensive, bulky, and pose an exposure risk above the maximum permissible exposure limit.[28, 55, 56]

Whether these changes in perfusion pose a significant risk to CKD/HD patients remains to be seen. A future longitudinal study predicting wound formation and correlating changes in perfusion with wound healing will help clinicians tailor therapeutic regimes to best serve each patient. For example, patients with poor perfusion can be provided advanced therapies such as hyperbaric oxygen therapy (HBOT) that are known to promote angiogenesis, perfusion, and wound healing.[57]

Changes in BP are also dependent on other confounding factors such as BMI,[58] diabetic status,[59] age[60] etc. A multivariate linear regression (**Fig. S7**) shows a strong positive correlation (R = 0.72) accounting for seven such confounding variables. Furthermore, the covariance matrix shows that the volume of UF removed has strong and positive covariance with the change in systolic pressure. Patients who had high volumes of UF removed, tended to show higher changes in pressure. In the healthy group (**Fig. S8**), the main contributing variable was the change in systolic BP (R = 0.81) whereas all the other confounding variables had relatively low covariance with each other. This means that the contributor to PA change was the change in systolic BP (p = 0.0019). Melanin, the molecule that gives skin its characteristic color is also a major absorber and source of PA signal.[61] Our group recently studied the effect of skin tone on PA oximetry.[61] In this work, we excluded any signals from the skin surface and since patients serve as their own controls between scans, and we only evaluated the change between scans; thus, skin tone should have minimal impacts. Furthermore, this study recruited a diverse set of CKD/HD patients and HD subjects from all possible skin types (Fitzpatrick scores 1-6) to account for skin tone bias. Other confounding variables could be changes in vascular physiology (increased resistance, calcification etc.) due to chronic diseased states in the CKD/HD groups,[62] but these are extremely difficult to account for.

### 4.1 Limitations

The LED-based PA system used in this work is an inexpensive, portable, and non-invasive imaging system. The LEDs used in this system operate 1000-fold under the maximum permissible exposure limit of 20 mJ/cm^2^.[63] But the system is limited in data processing as it condenses the number of frames between acquisition and export. Since we used a handheld scanner for all scans, it is difficult to generate 3D maps of the imaging area even though the system is capable to do so. The use of motion compensation and deep-learning algorithms to reduce noise and align frames could help map perfusion and oxygenation of the tissue.[64, 65] Future work will look to map tissue oxygenation in real time.

Within the CKD/HD group, hematocrit readings could help normalize PA intensity changes for variable fluid loss between patients. But not all dialyzers were equipped to measure hematocrit in real time limiting our analysis.

## 5. Conclusion

The management of BP and peripheral perfusion in dialysis patients is extremely important and challenging to monitor. Peripheral perfusion measurements are rare at the point-of-care. In this work, we imaged peripheral perfusion in 20 HD patients and 11 healthy subjects using PA imaging. We compared the peripheral tissue perfusion response to changes in blood pressure due to dialysis or exercise. The healthy group showed a positive correlation to changes in BP. The CKD/HD group also showed a positive correlation to BP changes during a dialysis session. In comparison, the CKD/HD group showed a significantly reduced PA response to changes in blood pressure compared to healthy controls.

## Data Availability

All data produced in the present study are available upon reasonable request to the authors

## Acknowledgments

**Fig. 1A-B** were created with BioRender.com. This work was supported by the National Institutes of Health through R21 AG065776 as well as internal funds from UCSD under the Galvanizing Engineering in Medicine program. YM and TD would like to acknowledge help from Yvette Waters, RN, BSN, CNN and the entire nursing team at the outpatient hemodialysis unit, UC San Diego Medical Center, Hillcrest, San Diego, CA, USA.

## Data Availability

The raw anonymized data that supports the findings of this study are available on reasonable request from the corresponding author.

## Supplementary material

Supplementary data associated with this article can be found in the online version.

## Abbreviations

CKD: Chronic kidney disease
ESKD: End stage kidney disease
HD: Hemodialysis
BP: Blood pressure
TcOM: Transcutaneous oxygen monitoring
fMRI: Functional magnetic resonance imaging
LDI: Laser doppler imaging

