## Supplementary material for "Monitoring peripheral hemodynamic response to changes in blood pressure via photoacoustic imaging": Table S1

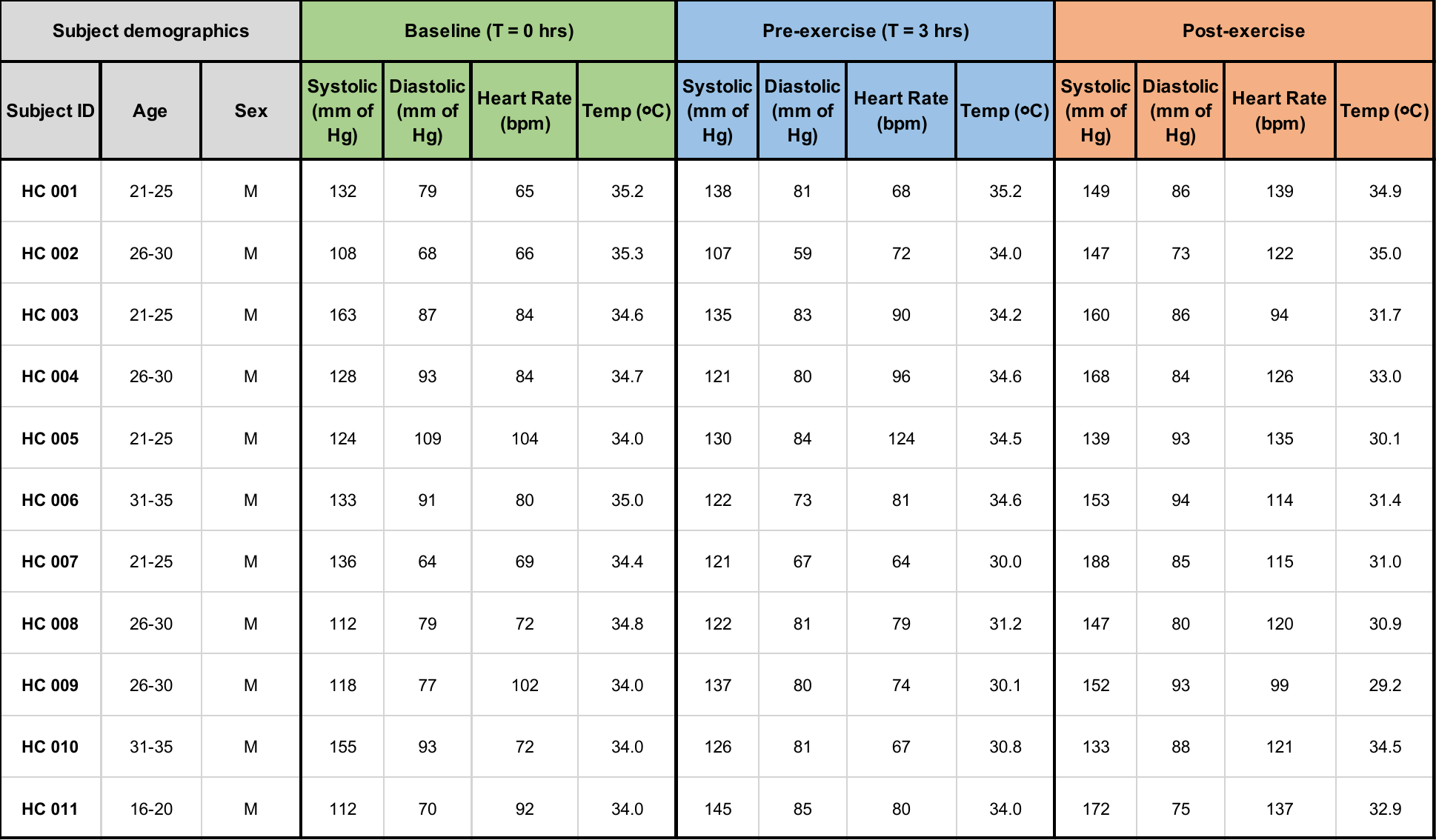

**Table S1. Healthy control group.** Blood pressure, heart rate, and temperature at baseline, pre-exercise, and post-exercise.

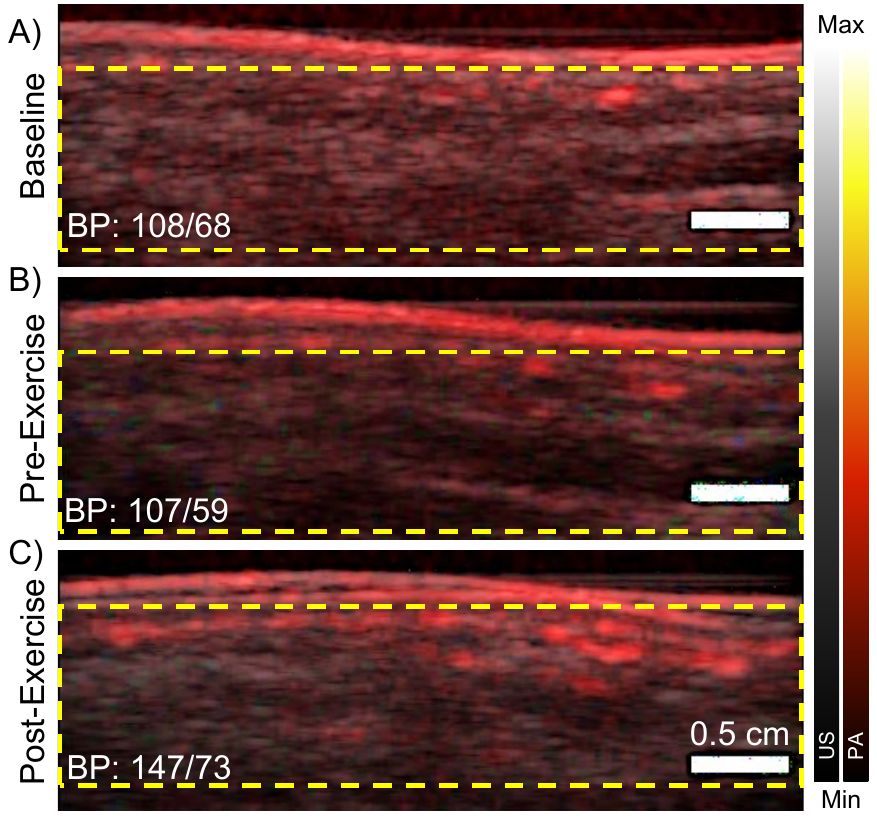

**Figure S1.** Region of interest analysis for quantifying PA intensity for healthy subjects (HC 002 in this figure). We used a 4 cm wide and 1 cm deep rectangular ROI that was kept constant for all scans. The skin layer was excluded from the analysis. Scale bars measure 0.5 cm. PA intensity and therefore perfusion increased with a rise in BP (40 mm of Hg increase after exercise). Scale bar represents 0.5 cm.

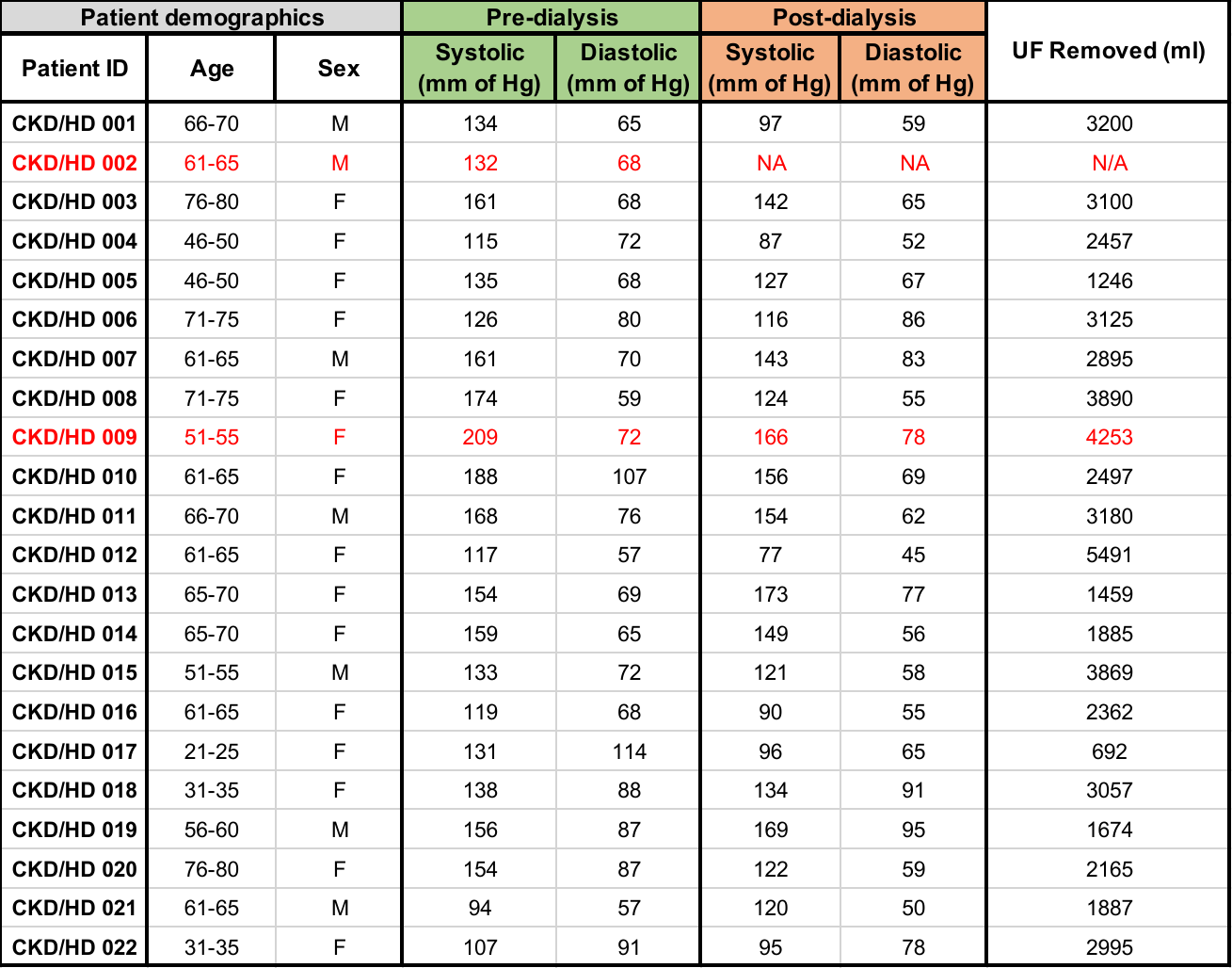

**Table S2. Diseased group.** 22 patients on hemodialysis were recruited for this study. We recorded blood pressure and ultrafiltrate removed (UF removed) before and after a dialysis session. Two patients (marked in red) were excluded from analysis due to complications independent of imaging during the session.

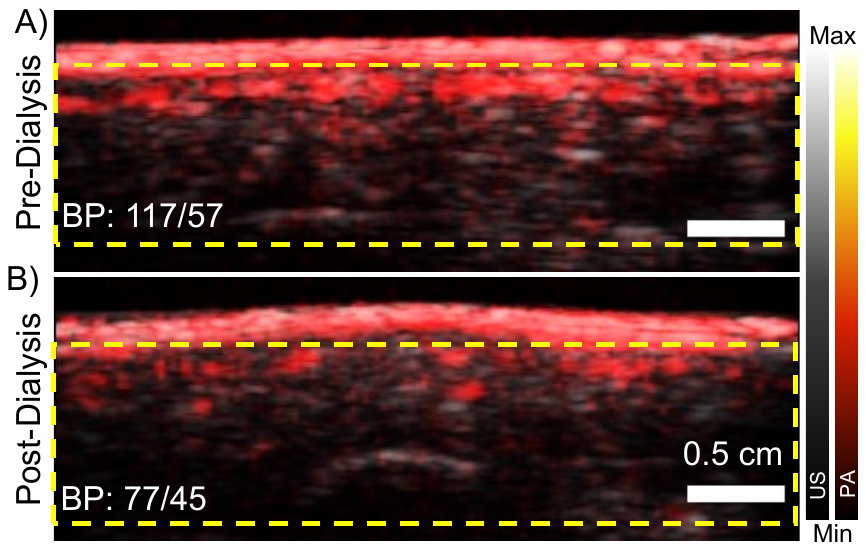

**Figure S2.** ROI analysis for the diseased group (CKD/HD 012 in this figure). These images are from the left heel. After dialysis (panel B), PG 012 experienced a 40-mm-Hg drop in BP resulting in reduced PA signal and perfusion. Scale bars represents 0.5 cm.

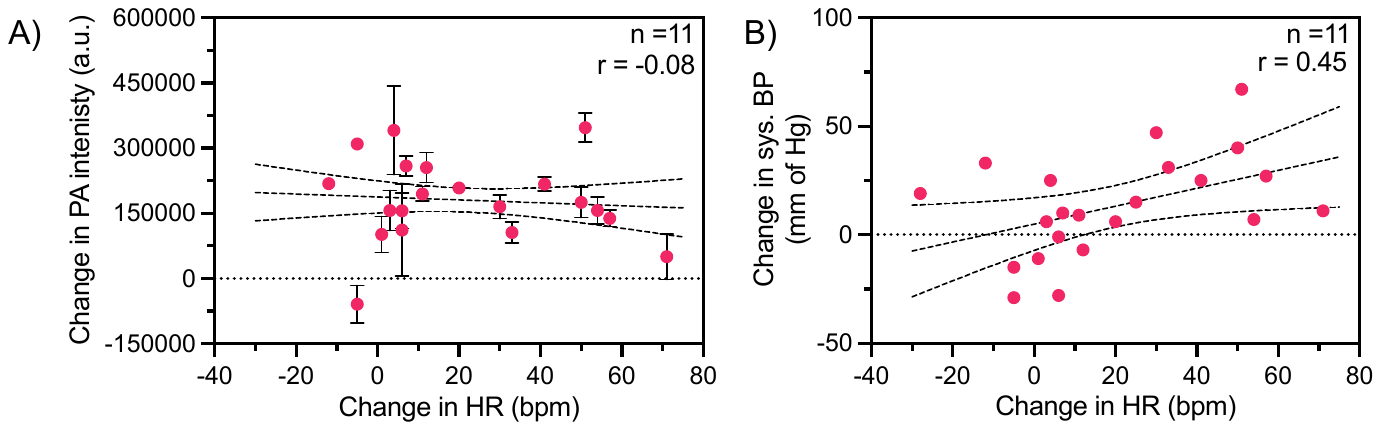

**Figure S3.** Effect of heart rate (HR) on perfusion and systolic BP in the healthy cohort. **A.** The change in HR had no significant effect on the change in PA intensity. **B.** The change in BP was positively correlated to an increasing HR due to exercise.

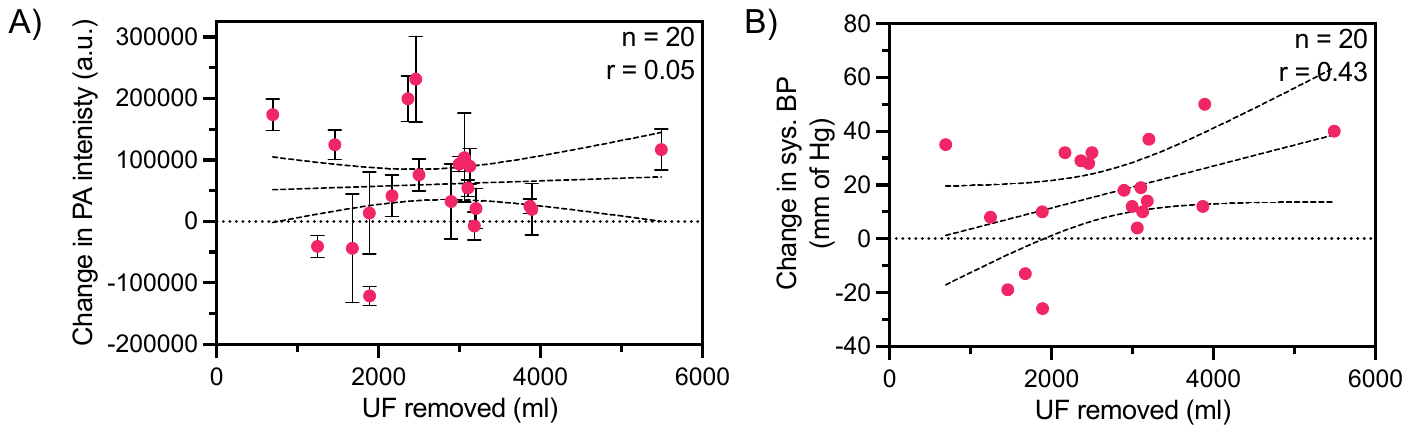

**Figure S4.** Effect of ultrafiltrate (UF) removed on perfusion and blood pressure. **A.** Changes in PA and perfusion were independent of the UF removed. **B.** The change in systolic BP was not significantly correlated to the amount of UF removed.

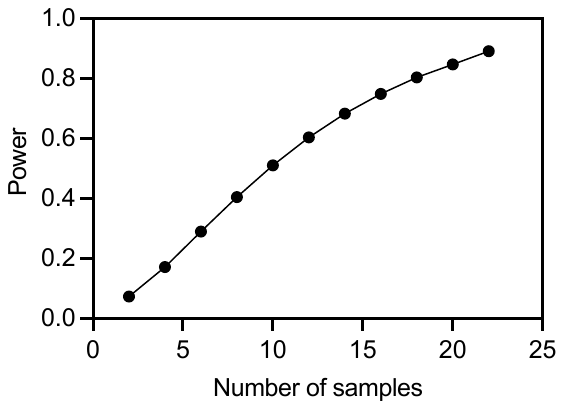

**Figure S5**. Power curve for the dialysis patients shows 85% power with n = 20.

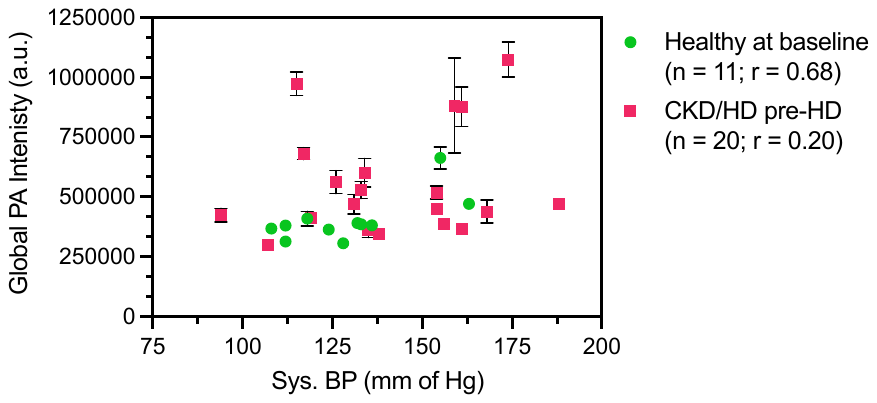

**Figure S6. Correlation between global PA intensity and absolute systolic BP at rest / baseline.** The healthy group shows a significant positive correlation (r = 0.68; p<0.05) compared to the CKD/HD group (r = 0.20, p>0.05).

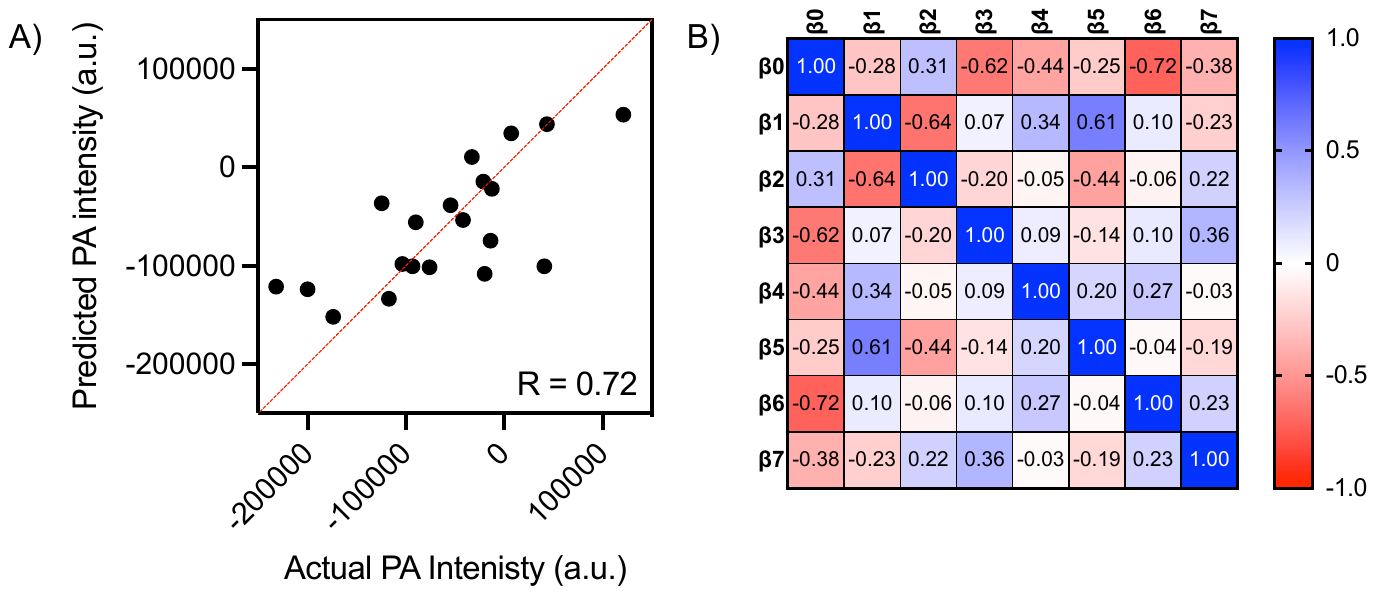

**Figure S7. Multivariate linear regression analysis for the CKD/HD group. A.** The coefficient of multiple correlation (R = 0.72) with 7 independent variables shows PA intensity can be predicted using the below-mentioned confounders. **B.** Covariance matrix reporting the degree of intertwining between the confounding variables. **Β0** = Intercept (mean response when all the variables = 0)**; β1** = change in Sys BP (mm of Hg); **β2** = change in Dia. BP (mm of Hg); **β3** = age (years); **β4** = sex (Female/Male); **β5** = UF removed (ml); **β6** = BMI (kg/m^2^); and **β7** = diabetic status (No/Yes). Higher positive values indicate strong covariance between those variables. For example, between **β5** and **β1** = 0.61 indicates that when a higher volume of UF is removed, the change in systolic pressure during an HD session also tends to be higher. Similarly, between **β7** and **β4** = -0.03 indicates that the patients BMI and sex are not related to each other.

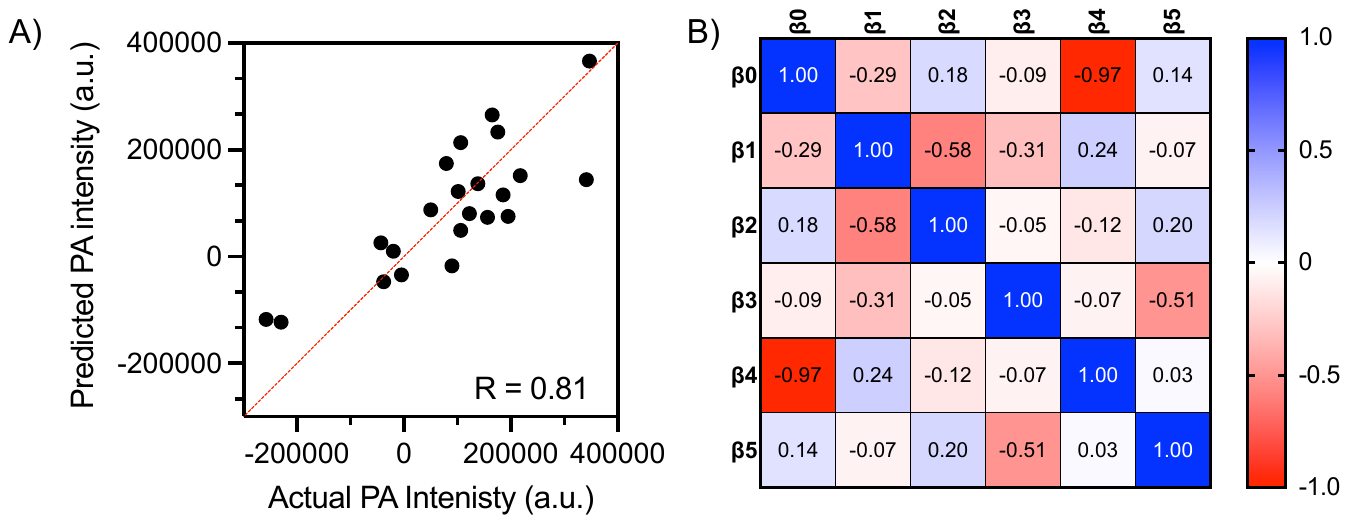

**Figure S8. Multivariate linear regression analysis for the healthy group. A.** The coefficient of multiple correlation (R = 0.81) with five independent variables shows PA intensity can be predicted using the below-mentioned confounders. **B.** Covariance matrix reporting the degree of intertwining between the confounding variables. **Β0** = Intercept (mean response when all the variables = 0)**; β1** = change in Sys BP (mm of Hg); **β2** = change in Dia. BP (mm of Hg); **β3** =change in heart rate (BPM); **β4** = Age (yrs); **β5** = change in body temperature (^o^C). Age showed a strong negative correlation with the intercept, meaning younger subjects showed higher PA signals. The covariance between all other parameters was low in the control group. This means that the main contributor to the correlation in panel A is the change in systolic BP (p = 0.0019).
